# Partial unlock model for COVID-19 or similar pandemic averts medical and economic disaster

**DOI:** 10.1101/2020.03.30.20048082

**Authors:** Robert L. Shuler

**Author notes:** Corresponding author (RS).

## Abstract

Data as of March 29, 2020 show that the “flattening” strategy for COVID-19 in the U.S. is working so well that a clean removal of social distancing (aka “unlock”) at any time in 2020 will produce a renewed catastrophe, overloading the healthcare system. Leaving the economy locked down for a long time is its own catastrophe. An SIR-type model with clear parameters suitable for public information, and both tracking and predictive capabilities which “learns” disease spread characteristics rapidly as policy changes, suggests that a solution to the problem is a partial unlock. Case load can be managed so as not to exceed critical resources such as ventilators, yet allow enough people to get sick that herd immunity develops and a full unlock can be achieved in as little as five weeks from beginning of implementation. The partial unlock could be for example 3 full working days per week. Given that not all areas or individuals will respond, and travel and public gatherings are still unlikely, the partial unlock might be 5 full working days per week. The model can be regionalized easily, and by expediting the resolution of the pandemic in the U.S. medical equipment and volunteers, many of them with already acquired immunity, can be made available to other countries.

## Introduction

After failure to contain COVID-19 in China, the global response has largely been social isolation, with effective cessation of global and local travel except by personal vehicle, and in some cities by bus and subway with frequent sterilization protocols. The strategy is called “curve flattening” and is intended to contain demand for special medical equipment, especially ventilators and related devices, from exceeding supply, which would cause unnecessary deaths.

A vaccine reputed to be 18 months away, the remaining strategy is herd immunity. For COVID-19 with an R0 (R-naught, number of people to whom each victim transmits the virus) of 1.4 to 3.28 [1], and seemingly everyone susceptible with no natural immunity, up to 70% of the population must have the disease for herd immunity to come into play. With half a million known and likely 2.5 million unknown cases worldwide, a high spread rate, many very mild cases, and a contagious incubation period, containment is no longer a viable option. The disease is substantially different than MERS or SARS. Although, many in the public and some officials seem to be behaving as if containment were viable.

In fighting a disease with public policy that relies on compliance, it is important to avoid the following sort of dilemma. Suppose we posit two policy choices, lockdown and unlock. Suppose the implementation of a policy has two parts: a planned scientific and administrative decision, and the public compliance with that decision. Suppose either one of those parts amounts to “If the disease is spreading and there is horror in the hospitals, lockdown, otherwise unlock.” Then lockdown reduces spread and produces a sense of relief, which results in unlock, which in turn restores the pandemic. A Gödel undecidability ensues and the actual strategy flip flops ineffectively. In this paper we propose a model which rapidly measures and uses public response to policy, rather than depending on how well a particular policy is implemented or accepted, dodging the dilemma. This leaves individuals free to self-isolate or risk working without overly compelling them. It leaves regional governments free to modify the policy to fit their area, if for example the disease is spreading faster and threatening hospital resources. To predict the effect of an “unlock” the model (or the person using it) can use a response calibration from a previous time, or estimate a new one. For this paper, we use the initial spread rate to model “unlock.” This may be high, as people will likely not go back to mass gatherings and flying on airplanes immediately, which simply means we model a worst case scenario which gives the model a margin of safety.

A principle objective of this paper is to share results and strategy possibilities before the conclusion of the COVID-19 outbreak. If studying frequently repeating phenomena, the scientific approach is to present complete data. However, we are studying a rare (roughly once in 100 years) phenomena, with the aim to intervene before it is concluded.

## Approach

The goal of our approach then discards containment as an opportunity past (and perhaps not realistic from early on), and a vaccine as a prospect too far in the future to avoid economic catastrophe. Opinions differ as to the effect of severe and prolonged recession on mortality and health. For example, there are fewer motorway deaths due to less driving [2]. On the other hand the 2008 financial crisis resulted over the next few years at least 260,000 additional cancer deaths [3]. Economic losses from pandemics, even without a long term global shutdown, have been estimated at the low end of but within the range of impacts from climate change [4].

Therefore the goal we adopt is to use public policy supported by modeling to achieve herd immunity as rapidly as possible, without overloading the healthcare system’s critical resources (we use ventilators as a general proxy for resources) and causing unnecessary deaths. Given our assumptions about vaccines, containment and R0, 70% of the population will get sick anyway. Our approach of voluntary compliance with unlocks allows individuals and regional governments to manage their health and economic risks as they see fit. As the U.S. has ventilators per capita among the highest in the world, and stands to suffer high economic damage, it is a logical place to attempt this strategy. If the U.S. develops herd immunity, then it might choose to make some of its ventilators, and its production capacity for more ventilators, available to other countries. At the current time the hoarding mentality which arises from uncertainty is impairing the sharing of medical equipment and New York and Italy plead for ventilators and masks, as other countries protest they need their own stockpiles. But China, feeling it has the spread under control, is more forthcoming.

### Model parameters

The purpose of this paper is more to illustrate an approach than to propose a specific model. We suggest following Huppert and Katriel’s guidance: “*To examine which of the predictions made by a model are trustworthy, it is essential to examine the outcomes of different models. Thus, if a highly simplified model makes a prediction, and if the same or a very similar prediction is made by a more elaborate model that includes some mechanisms or details that the first model did not, then we gain some confidence that the prediction is robust. An important benefit derived from mathematical modelling activity is that it demands transparency and accuracy regarding our assumptions, thus enabling us to test our understanding of the disease epidemiology by comparing model results and observed patterns. Models can also assist in decision-making by making projections regarding important issues such as intervention-induced changes in the spread of disease*.” [5]

To that end we select parameters according to the following criteria:

1. Provide sufficient realism to assure relevance
2. Keep the model simple
3. Compatible with easily accessible public data for updating/tracking
4. Err toward the worst case rather than optimism

We expect numerous models to come into play in an actual policy implementation, and regional authorities to do their own modeling. In addition, we want our model to be comprehensible by the public and to aid in the important goal of informing the public and enlisting their rational choices in implementing the strategy. To this end our first parameter reformulates the traditional R0 reproduction rate, which does not have a specific time base, as a time-based spreading rate “R.” Data from the CDC https://www.cdc.gov/coronavirus/2019-ncov/cases-updates/cases-in-us.html#investigation for number of cases for March 16-21 in the US (4226, 7038, 10442, 15219, 18747, 24583) represent a time period when local spread had begun to act exponentially. These data correspond to daily spread rates of (1.665, 1.48, 1.45, 1.23, 1.311). Previously a slow rate attributable to travel had been dominant. After March 21 the rate of spread decreases continuously, which we presume is due to increased government directed social distancing including widespread business closure, stay at home orders, and increased public compliance with these policies. For the model’s initial value we adopted a daily spread rate of 1.414. The fraction of *new* cases is then R=0.414. This number is immediately replaced in the model by the next day’s rate, and adjusted by the computed herd immunity factor when re-used following unlock. The unlock spread rate should be measured and adjusted, but there has yet been no unlock.

Our R can be compared with R0 by selecting a time window for transmission. Several possibilities exist, and since our R is empirically measured we are not too concerned with determining the exact relation. But for example, suppose there is a window of 4 days before a person discovers they have the disease and self-isolates. At our presumed R (cases doubling every two days) there would be 4 people with the disease at the end of this time, or 3 new cases. That corresponds to R0=3, which is in line with the high end of estimates given above (3.28 maximum).

The number of ventilators in the U.S. including reserves, alternatives (anesthesia machines) and older equipment is taken at 200,000 [6].

The ratio of total likely cases to known reported cases is taken at 6 (16.6% known) with alternative scenarios checked at 5 (20% known). The lower numbers are more critical due to the way the model calculates ventilator requirements. Published numbers are typically around 14% [7], so our assumption has some margin of safety.

A precise number for how long a case of COVID-19 lasts is of course not obtainable due to the wide variation. However this number is not critical for our model. They could last forever and the herd immunity calculation would be unaffected since it includes both recovered and infected people. Somewhat more important is the length of time a ventilator is required, if one is required. The best estimate we could obtain was “up to weeks” since this too is highly variable. We used 14 days for both numbers.

The fraction of cases which require resources such as a ventilator is also important. We used 3.2% of known cases, or about half of critical cases, taken from Meng, et. al. [8] Lower estimates exist. This is an area in which additional data and input from more specialized simulations would be used before public policy formulated. Regionalization is also important as ventilators may not be distributed where needed. Publication daily of model predictions, assuming they predict or exceed the actual data coming in, we believe would increase confidence and promote redistribution of ventilators according to need.

Social networks, location tracking and other massive data mining efforts recommended in research of more persistent (non-pandemic) diseases [9] are specifically not part of our approach. They take time, where we require rapid feedback. They invite abuse for other applications later. But most important, social networks change as soon as a pandemic is announced, change again when government policy is announced, and keep changing. An aggregated tracking and feedback method will work better.

### Model dynamics

We use a standard SIR modeling approach [10] but with the time based reproductive factor as described above. The number of new cases is our R factor times the existing cases which are only 4 days old or less.

During “lockdown” the R factor is adjusted according to (a) the ratio of new cases from the previous day, and (b) the increase in herd immunity factor over the previous day. Only when the caseload is rapidly peaking will day to day changes in herd immunity factor be relevant. When an “unlock” policy is established in the predictive model, the R factor is re-established at the initial value reduced by the herd immunity factor. When actual unlock data is available, this will need further adjustment which we do not specify at this time.

This simple model was implemented in a spreadsheet with some facilities for setting days to be unlocked, or they can be set manually. The spreadsheet will be available as supplementary materials and in the http://medRxiv.org preprint repository, and is currently available at http://shulerresearch.org/covid19.htm. Each day actual data was used to replace predicted data based on CDC data at the link specified above. This affects the model’s integration base and the effective reproduction rate R. The number of “new” cases, necessary for bookkeeping active and spreading cases and ventilator utilization, is deduced from the day to day change is number of cases. Total cases, used for the herd immunity calculation, is calculated by either 5 or 6 times the known cases as described.

## Results

The progress toward “curve flattening” is shown in Figure 1, with predicted plots based on actual data from 3/21, 3/25 and 3/29. One can see that to say flattening was successful is putting it mildly. Thus arises the dilemma: not enough people are getting sick to develop herd immunity.

**Figure 1.**
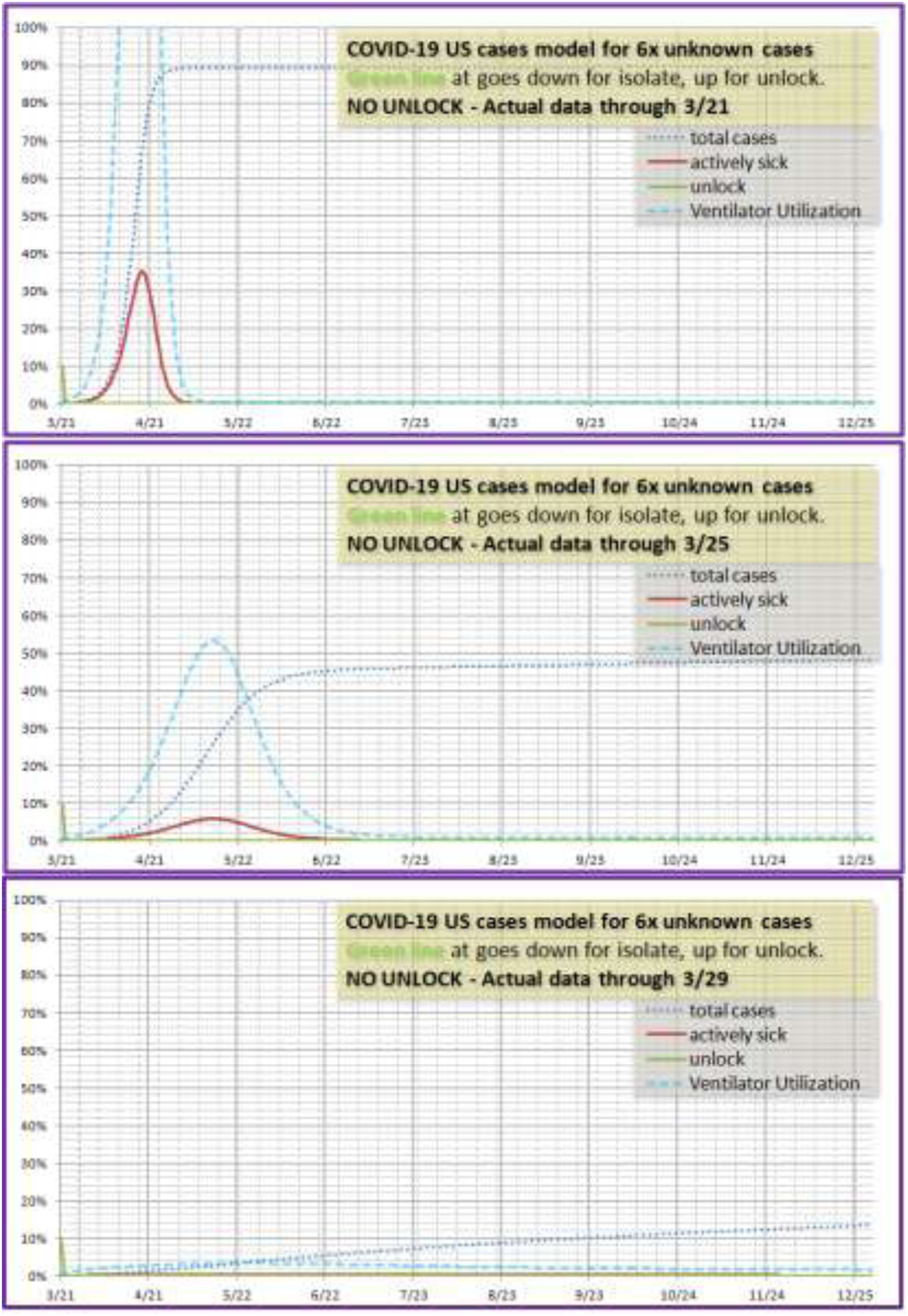
Progress toward “flattening” from 3/21 to 3/29.

In the 3/25 and 3/29 cases in Figure 1, herd immunity is incomplete. In the 3/21 case it appears 90% of the population is infected because of numerical overshoot during the rapid rise. One day quantization may not be enough resolution, the rise is so rapid. The extent to which such an overshoot might happen in reality (it might) is not of concern to us since this case will not be allowed. In the following cases it will be apparent that where a slower rise is allowed due to partial lockdown, the total cases peaks around 70% just where it should for R0=3.

Figure 2 shows two different dates for total unlock, beginning with the day after the current extension of lockdown through April 30^th^. The second scenario begins 6 months later on November 1. Though the peak is reduced somewhat, ventilator capacity is still wildly exceeded. More ventilators may be produced by then, but 6 months have been wasted when people could have been building herd immunity, and the economic loss, the permanent loss of houses and business from loan default, and consequent loss of jobs, will be known is the worst “man-made” disaster of all time, since it was a reactionary policy decision which we will show is easily prevented.

**Figure 2.**
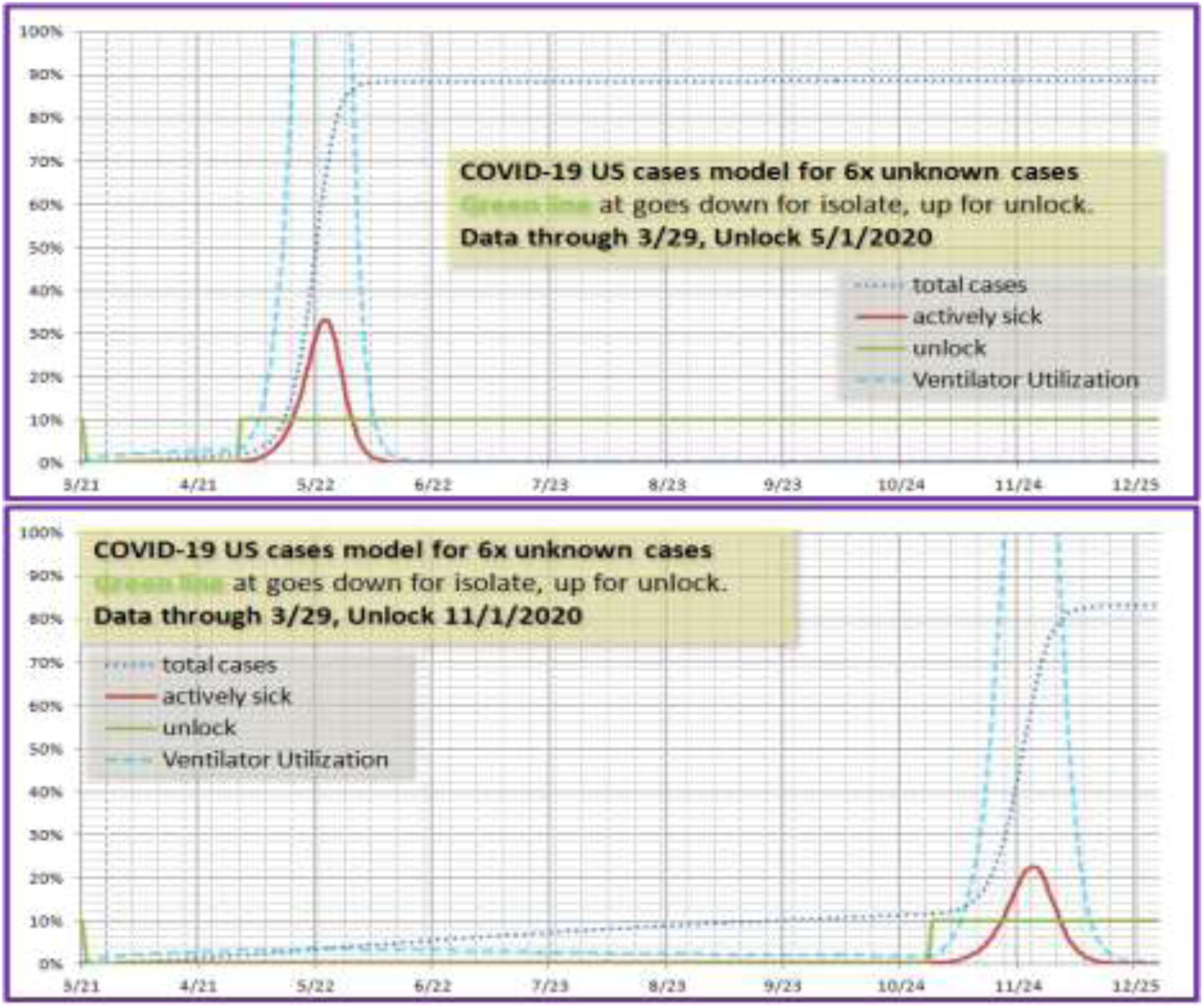
Alternatives for full unlock.

Figure 3 shows a three day a week partial unlock strategy beginning as soon as April 9^th^. Actually it could begin April 2^nd^, but this paper is not likely to be in public view by then. Five weeks later, total unlock is possible. Critical resource (ventilator) usage expands smoothly toward the 70-80% range, then drops dramatically just at the time of full unlock. It is possible to unlock a week earlier, but doing so causes a rise in critical cases which might undermine public confidence.

**Figure 3.**
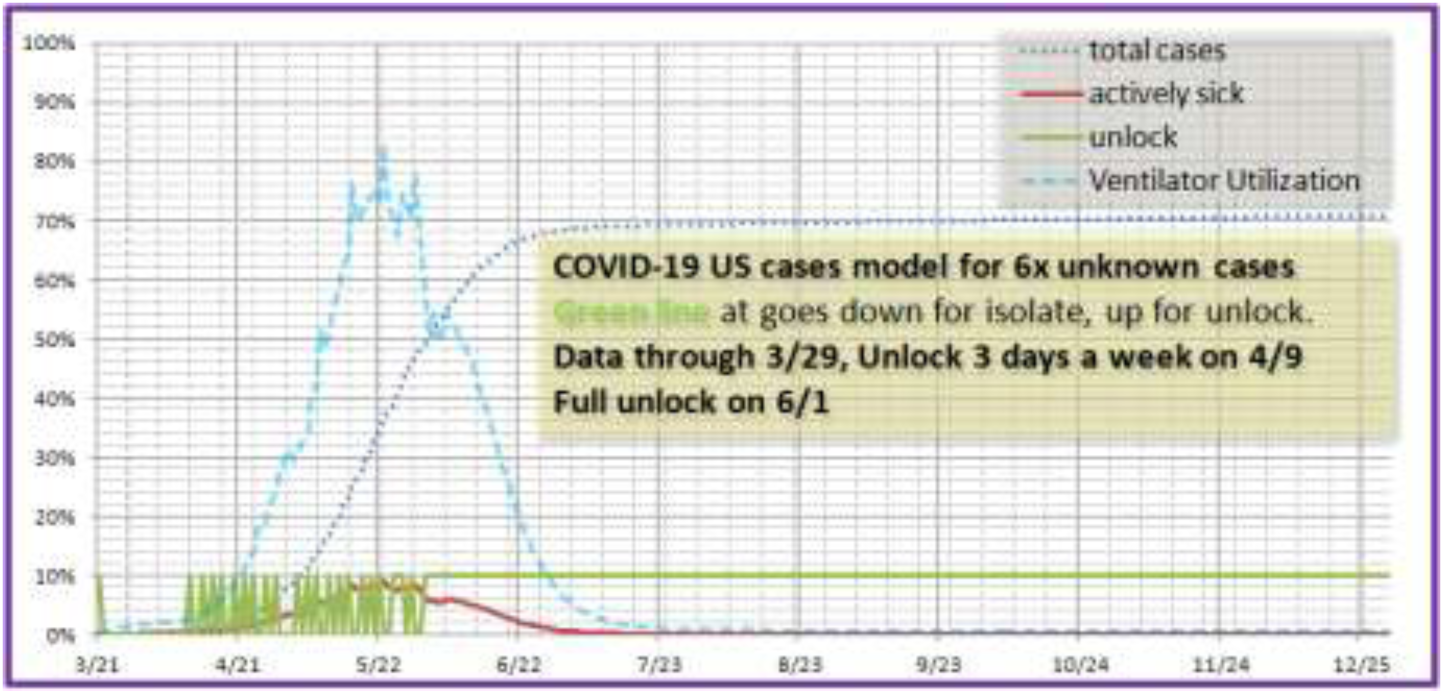
Three day a week unlock scenario.

Figure 4 shows a sensitivity analysis for the multiple of unknown to known cases of 5 instead of 6. The situation is worse, but capacity is not exceeded. Most expert opinion is that the multiple will be higher rather than lower, however additional studies involving testing random samples would clarify this figure, which might vary by region.

**Figure 4.**
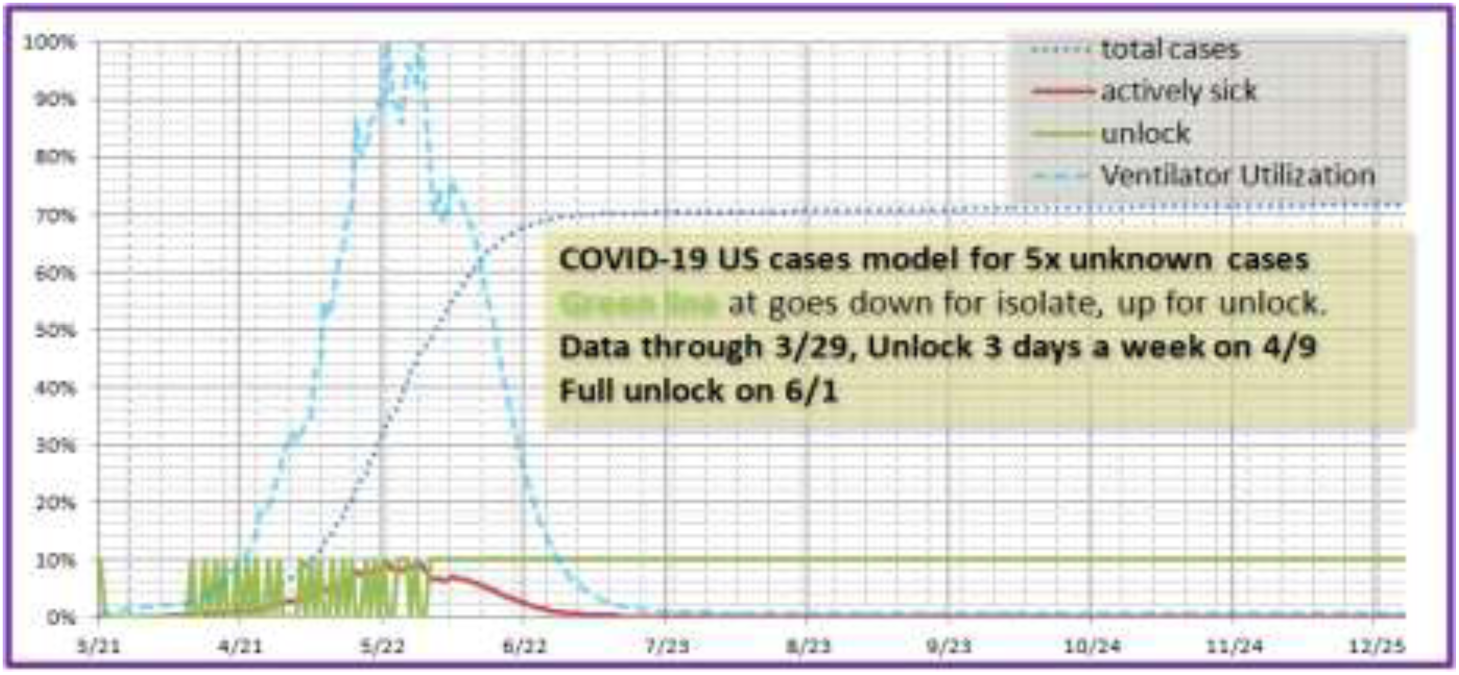
Three day a week unlock scenario with 5x unknown to known cases.

It appears that the unlock date need not be pinned down in advance. There will be a precipitous drop in usage of critical resources, and unlock can then occur within a few days.

## Conclusion

It appears that options are available that are not now openly under consideration that would lessen economic impact of the CORVID-19 worldwide response by six to ten times, while operating within existing critical medical resources and not causing unnecessary deaths.

The public desperately wants more concrete data and projections, projections which can be seen to be valid as the data comes in. Current models involve obscure parameters that make investigators hesitant to share them. The use of a time base reproductive factor would likely be more comprehensible and easier for the public to verify for themselves.

It is likely the response to a “partial unlock” would be underwhelming. Many businesses would be hesitant to reopen. However, within a few days this effect will be apparent in the case data, and the unlock schedule can be adjusted higher or lower. Using days of the week is a customary and usual way of managing work schedules that is easily understood.

The public will realize, and it must be made clear, that partial unlock is not a declaration of safety from CORVID-19. It is a declaration of reasonable assurance of adequate medical capacity. We recommend a flexible policy allowing those at risk or those simply afraid to continue to isolate, and those who wish or need to work to accept the risk. The end result is a great reduction of the isolation time required for those who choose to continue to isolate.

The final and perhaps most important benefit of this methodology and strategy is the rapid availability of excess medical resources from the U.S. (once unlock is achieved, in about 5 weeks from start) to assist the rest of the world which is not so fortunate in number of ventilators or comparable devices. Further, healthcare workers who have acquired immunity may well want to volunteer to help in other countries. Let’s do this.

## Data Availability

The model file will be uploaded as a supplementary file. It is also available at shulerresearch.org. The small amount of data is available from the CDC at a link provided in the paper.

http://shulerresearch.org/covid19.htm

## Acknowledgements

The author appreciates feedback, comments, references and general moral support from Theodore Koukouvitis at every stage of this research, and for some time previously as the author expanded his modeling and theoretical work in social systems safety and reliability into cooperative and evolutionary/biological systems.

